# Economic costing of evaluating, deploying and monitoring an artificial intelligence-based reconstruction for acceleration of rectal MRI examinations

**DOI:** 10.64898/2026.05.18.26353474

**Authors:** Ciara A Harrison, Mengjun Wu, Owen A White, Georgina Hopkinson, Julie Hughes, Scott Robertson, Erica Scurr, Joshua Shur, Francesca Castagnoli, Geoff Charles-Edwards, Dow-Mu Koh, Jessica M Winfield

**Author notes:** Ciara Harrison and Mengjun Wu contributed equally to this work.

## Abstract

**Objectives:** AI-based reconstructions can reduce MRI acquisition times and/or improve image quality. Guidelines recommend clinical evaluations and post-deployment monitoring of these novel methods, however, there has been little investigation of the clinical resources required for such assessments. The aim of this study was to evaluate the healthcare resource utilisation and potential savings associated with AI-based reconstructions in rectal MRI.

**Methods:** A retrospective economic costing analysis was conducted from the NHS healthcare perspective. Resource utilisation data were extracted from the Electronic Patient Records for 9 healthy volunteer scans and 104 rectal MRI examinations evaluating an AI-based reconstruction. The resource profile included the MRI scan and the staff time required for data acquisition and analysis.

**Results:** The clinical evaluation of the AI-based reconstruction cost £15,023. Deployment of the AI-based reconstruction reduced the length of an MRI rectum scan by 22 minutes, theoretically saving approximately £3,437 per month. Addition of post-deployment quality control scans reduced this monthly saving to £2,636. If the quality control scans were evaluated using radiologists rather than image quality metrics, monthly savings would be approximately £2,541. With ongoing quality control, the clinical evaluation cost would be recouped between 5.8 and 6 months, compared with 4.4 months without ongoing quality control.

**Conclusions:** Deploying AI-based reconstructions can yield cost savings through reduced scanning times. Quality control tests using image quality metrics would save radiological burden and reduce costs compared with conducting repeated image scoring by radiologists.

**Advances in knowledge:** This study evaluates the healthcare resource utilisation and potential cost savings from implementing AI-based reconstructions in rectal MRI.

## Introduction

AI-based reconstructions can be used to accelerate and/or improve the image quality of MRI scans. These acceleration techniques have been proposed as one method to increase the throughput of patients through radiology services which are experiencing increasing demand^1^.

National and international guidelines recommend pre-deployment clinical evaluations of AI tools in radiology^2–6^. AI-based reconstructions may require thorough optimisation to establish a protocol to maintain image quality whilst achieving substantial time savings. A clinical evaluation should be performed in the relevant patient populations and measured relative to an agreed standard. Our institution previously carried out such an optimisation and evaluation of an AI-based reconstruction method^8^ in rectal MRI, reducing the total acquisition time by over 50% whilst maintaining or improving image quality^7^.

Guidelines also recommend that AI-based tools in imaging should be monitored due to known issues around performance drift, sensitivity to variation in input data, hallucinations, sensitivity to hardware degradation or changes due to scanner software upgrades^8–11^. While there are no specific recommended methodologies for monitoring AI-based reconstruction tools, radiology departments can adopt a routine quality control (QC) framework that aligns with established QC measurement principles: resource-efficient, quantitative, objective, reproducible, simple to perform and sensitive. However, special considerations for AI-based reconstructions must be made as conventional methods may not be appropriate: phantom-based methods have not been validated for assessing AI-based reconstruction methods trained on human data and radiologist assessments are resource-intensive. We developed a QC programme using quantitative image quality metrics (IQMs) that have been shown to be sensitive to known perturbations in the AI-based reconstruction as a resource-efficient alternative^12^.

AI tools are often presented in business cases as reducing costs^13,14^. In the specific application of AI-based reconstructions to accelerate scans, savings can theoretically be achieved by using AI to reduce the duration of an MRI examination, therefore enabling more examinations to be carried out without an associated increase in radiographer workforce requirements. However, the guidelines have little consideration of the clinical resources such as personnel and equipment required to evaluate and monitor AI-based reconstructions. The costs of such resource use must be assessed to fully evaluate the actual savings. Guidelines recommend that users should consider cost-effectiveness of AI tools in medical imaging ^2–4^, but economic costings are rarely performed in clinical practice ^15–17^.

The aim of this study was to evaluate the healthcare resource utilisation in evaluation and monitoring of AI-based reconstructions in rectal MRI examinations and potential savings for one specialist NHS trust, versus standard imaging.

## Methods

This study retrospectively evaluated the costs of three phases of implementing an AI-based reconstruction: clinical evaluation, clinical use, and ongoing monitoring. The imaging data in patients and healthy volunteers were acquired as part of a prospective study approved by a national research ethics committee (ClinicalTrials.gov Identifier: NCT05118555). All patients gave verbal consent for the additional research sequences as part of their clinical examinations. All healthy volunteers gave written consent.

Two 1.5 T systems and two 3 T systems were included in this study (MAGNETOM Sola/Vida, Siemens Healthineers, Forchheim, Germany). Subjects were scanned using a 30-channel anterior body array (or 18-channel anterior body array for some volunteer examinations) and a 32-channel posterior spine array. The AI-based reconstruction methods used in this study were MR scanner manufacturer products (Deep Resolve, Siemens Healthineers, Forchheim, Germany)^18,19^.

### Clinical evaluation and deployment

Initial protocol optimisation using 8 healthy volunteers was performed across 9 scanning sessions between January 2023 and March 2023. The AI-based reconstruction methods were applied in two sequences: a T2-weighted (T2w) sagittal turbo spin-echo (TSE) and a small field-of-view (sFOV) T2w axial TSE for rectal imaging. The sequences demonstrating fastest acquisition time whilst maintaining acceptable performance, from a qualitative review by radiologists, physicists, and radiographers, were selected for use in prospective patient imaging.

Fifty patients referred for a routine anorectal MRI between April 2023 and October 2023 underwent a single MR examination on either a 1.5 T (n = 22/50) or 3T (n = 28/50) scanner. The AI-based reconstruction was applied in accelerated sagittal and sFOV axial T2w-TSE sequences that were acquired in addition to standard-of-care T2w-TSE imaging at matched slice positions. Two radiologists visually assessed the paired sagittal and axial images and scored them using a 4-point Likert scale. Interim analysis of the radiologists’ scores after 20 patients indicated the image quality of the accelerated sequences at least matched that of the standard sequences. The accelerated sequences were deployed into the clinical protocol, replacing the standard sequences. The remaining 30 patients were scanned using the accelerated protocol with additional standard imaging at matched slice positions to acquire a reference dataset. Full details of the volunteer and patient evaluation can be found in previous work^7^.

### Ongoing monitoring

Between May 2024 and July 2025, additional QC data were acquired on one rectum examination per scanner per month. The additional acquisition was the standard sFOV axial T2w-TSE sequence at a matched slice position to an accelerated sequence in patients undergoing an accelerated rectum scan. The duration of the clinical evaluation, clinical use and monitoring phases are shown in Figure 1.

**Figure 1.**
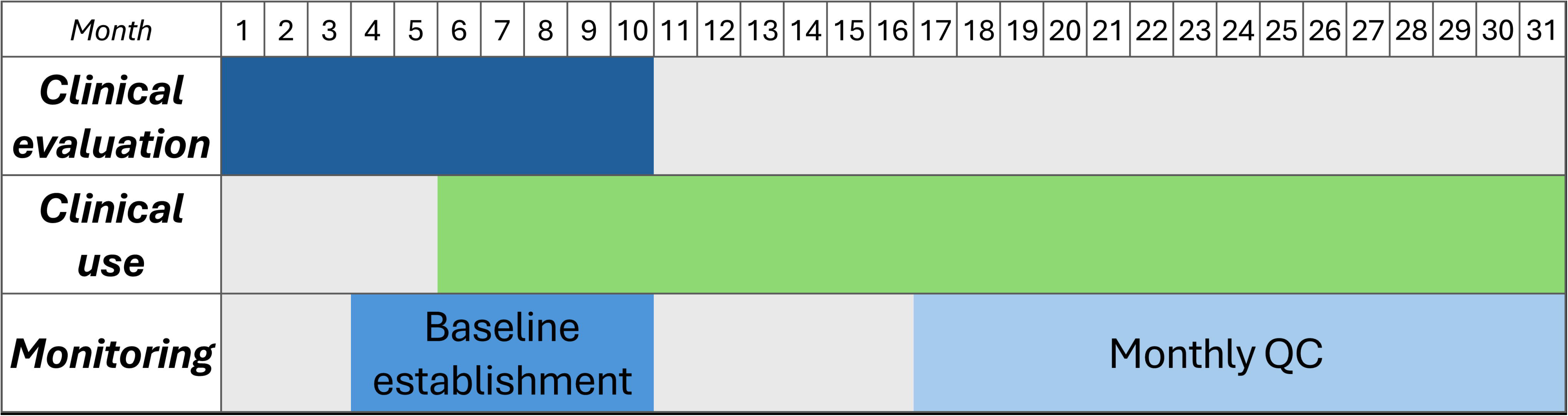
A Gantt chart indicating the duration of the clinical evaluation, clinical use, and monitoring shown up to July 2025. AI-based sequences remain in clinical use and monitoring is ongoing at the time of writing.

The image data and raw data were saved after each QC scan. A range of IQMs were evaluated on the image data, as described previously^12^. The IQM values from the first 50 examinations were used to establish baseline control limits, the IQM values from the QC examinations were used to perform ongoing monitoring. A band 8a physicist was involved in the first 8 QC scans, a band 7 physicist was involved thereafter. Three potential monitoring schemes were considered: no QC, QC using IQMs, QC using radiologists’ assessment of image quality.

### Health Economics

Economic costing was conducted from a healthcare perspective by a health economist. Resource use data were extracted retrospectively from the Electronic Patient Record of 8 healthy volunteers and 50 rectal patients who were scanned for the clinical evaluation. Given that resources used in this study were identified at very detailed level and the resource use data were collected at each patient level, this study employed a bottom-up micro-costing methodology^20^.

A detailed resource profile consisted of estimated time of work undertaken by involved staff members: administrative staff, radiology department assistants, physicists, radiographers, radiologists and referrers; and equipment resources: MRI scans and antispasmodic injections. Times of work were either calculated via lengths of examinations on the radiology information system (RIS), via lists on the picture archiving and communication system (PACS), as an average per examination, or estimated by the member of staff as the average time taken to do the task.

For each type of healthcare resource use, an appropriate unit cost was identified and valued using the NHS reference costs^21^, Personal Social Services Research Unit resources^22^ or the NHSBSA Drug Tariff^23^. The value of each resource use (ie, the quantity of resources affected multiplied by the unit cost) was estimated, then a total cost profile was attached to each examination.

### Costing resource use

Table 1 presents unit costs applied to resource use data. The unit cost of each MRI rectum scan was £156.05^21^. The costs of the staff members involved with the examinations ranged from £0.38 to £2.38 per minute. Most patients received an injection of hyoscine butyl bromide 20mg (Buscopan) intramuscularly before the scan, while a minority received glucagon (a small number received neither due to contraindications). The incurred cost was £0.40 or £11.52 for each injection of Buscopan or glucagon, respectively.

**Table 1.**
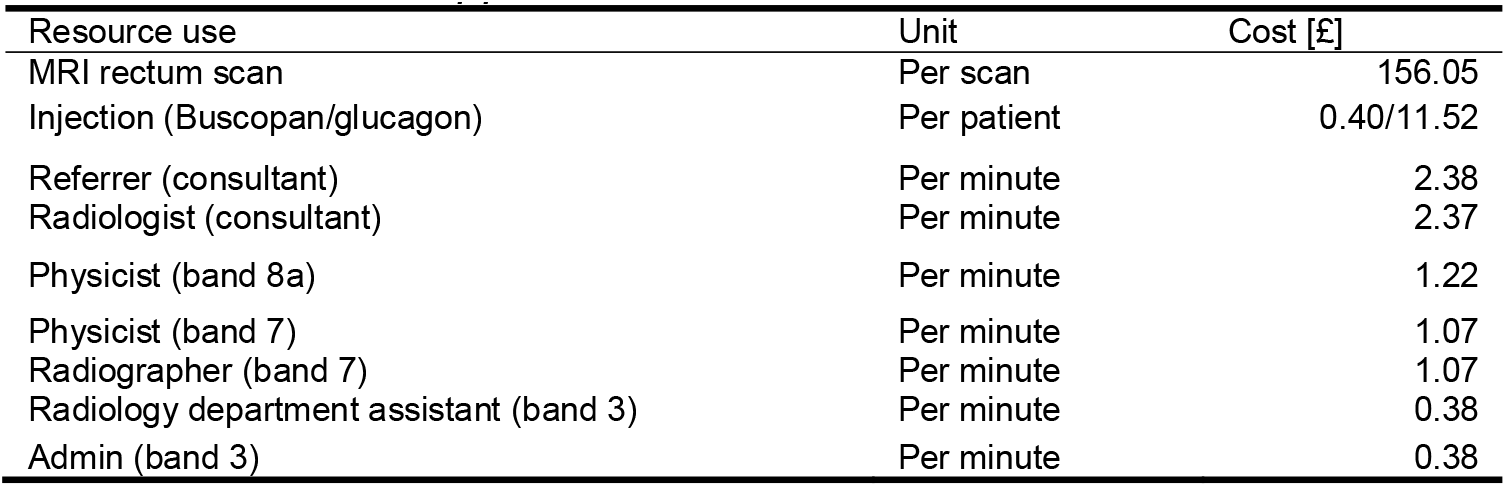
Unit costs applied to resource use data.

| Resource use | Unit | Cost [£] |
| --- | --- | --- |
| MRI rectum scan | Per scan | 156.05 |
| Injection (Buscopan/glucagon) | Per patient | 0.40/11.52 |
| Referrer (consultant) | Per minute | 2.38 |
| Radiologist (consultant) | Per minute | 2.37 |
| Physicist (band 8a) | Per minute | 1.22 |
| Physicist (band 7) | Per minute | 1.07 |
| Radiographer (band 7) | Per minute | 1.07 |
| Radiology department assistant (band 3) | Per minute | 0.38 |
| Admin (band 3) | Per minute | 0.38 |

The payback period was defined as the time between deploying the AI-based reconstruction and the time point at which the savings from accelerating examinations and performing a given monitoring scheme would be equivalent to the costs of performing the initial clinical evaluation.

## Results

### Volunteer study costing

Table 2 presents the staff costs for each volunteer scan. The data acquisition by the radiographers was 60 minutes, which cost £64.20 per scan, the equivalent task for the physicist was the costliest portion of the volunteer scan, costing £73.20 per scan. The total costs of staff use for the volunteer study was £3,042.23.

**Table 2.** Staff use for each volunteer scans and costs per scan.

| Staff | Tasks | Time per scan [minutes] | Costs per scan [£] |
| --- | --- | --- | --- |
| Physicist<br>(band 8a) | Recruitment & scheduling | 20 | 24.40 |
|  | Set up imaging protocol on scanner | 30 | 36.60 |
|  | Data acquisition | 60 | 73.20 |
|  | Data transfer & archiving | 30 | 36.60 |
|  | Data analysis | 56.25 | 68.63 |
| Radiographer | Vetting request | 5 | 5.35 |
|  | Data acquisition | 60 | 64.20 |
|  | Post-scan activity | 5 | 5.35 |
| Radiologist | Reviewing images | 10 | 23.70 |
| Total staff costs |  |  | 338.03 |

### Patient study costing

Table 3 presents resource utilisation and total average costs per patient scan for the clinical evaluation phase using standard and AI-based reconstructions. Data acquisition was performed by two radiographers: 50 minutes per radiographer for the first 20 patients (£107.00 per scan) and 35 minutes for the last 30 patients (£74.90 per scan). The same timings apply to the single physicist involved. The reduction in acquisition time reduced the total staff costs from £510.40 to £460.00. The average cost of injection for each scan was £1.24 (40/50 had Buscopan, 4/50 had glucagon, 6/50 had neither). Multidisciplinary team (MDT) preparation was 8.5 minutes on average given that 17/50 patients had imaging reviewed at MDT and it took an approximately 25 minutes per patient to prepare the imaging. The total average resource cost per scan using the 35-minute protocol was £617.29.

**Table 3.**
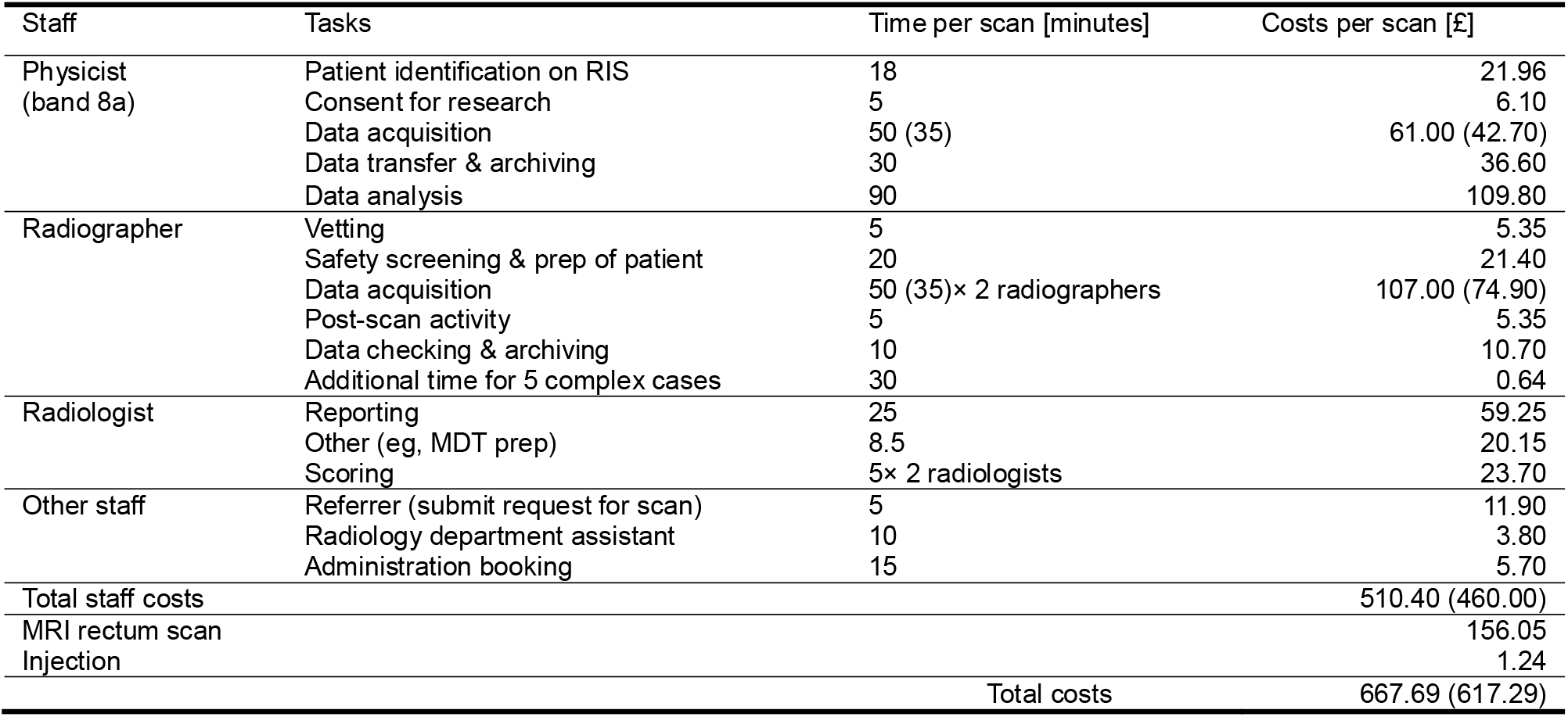
Resource use and costs per scan for the clinical evaluation phase where images were acquired using both standard and AI-based reconstructions. Values in brackets indicate times and costs for the last 30 patients.

Table 4 presents the resource use and total average costs on resource use per scan for the clinical protocol using AI-based reconstruction versus standard imaging. With standard imaging, the data acquisition time was 45 minutes on average and the total average cost of resource use per scan was £397.83. With the AI-based reconstruction, the data acquisition time was reduced to 23 minutes on average and the total average cost of resource use per scan was £350.75. Examples of imaging from the standard protocol and accelerated AI-based protocol are shown in Figure 2.

**Table 4.** Resource use for each patient and costs per scan using for the final clinical protocol using AI-based reconstruction and the original imaging protocol using standard imaging only. Rows in bold show values that differ between the two imaging protocols.

| Staff | Tasks | AI-based reconstruction |  | Standard imaging |  |
| --- | --- | --- | --- | --- | --- |
|  |  | Time per scan [minutes] | Costs per scan [£] | Time per scan [minutes] | Costs per scan [£] |
| Radiographer | Vetting | 5 | 5.35 | 5 | 5.35 |
|  | Safety screening & prep of patient | 20 | 21.40 | 20 | 21.40 |
|  | Data acquisition | **23 × 2 radiographers** | **49.22** | **45 × 2 radiographers** | **96.30** |
|  | Post-scan activity | 5 | 5.35 | 5 | 5.35 |
|  | Data checking & archiving | 10 | 10.70 | 10 | 10.70 |
|  | Additional time for 5 complex cases | 30 | 0.64 | 30 | 0.64 |
| Radiologist | Reporting | 25 | 59.25 | 25 | 59.25 |
|  | Other (eg, MDT prep) | 8.5 | 20.15 | 8.5 | 20.15 |
|  | Referrer (submit request for scan) | 5 | 11.90 | 5 | 11.90 |
| Other staff | Radiology department assistant | 10 | 3.80 | 10 | 3.80 |
|  | Administration booking | 15 | 5.70 | 15 | 5.70 |
| **Total staff costs** |  |  | **193.46** |  | **240.54** |
| MRI rectum scan |  |  | 156.05 |  | 156.05 |
| Injection |  |  | 1.24 |  | 1.24 |
| **Total costs** |  |  | **350.75** | **Total costs** | **397.83** |

**Figure 2.**
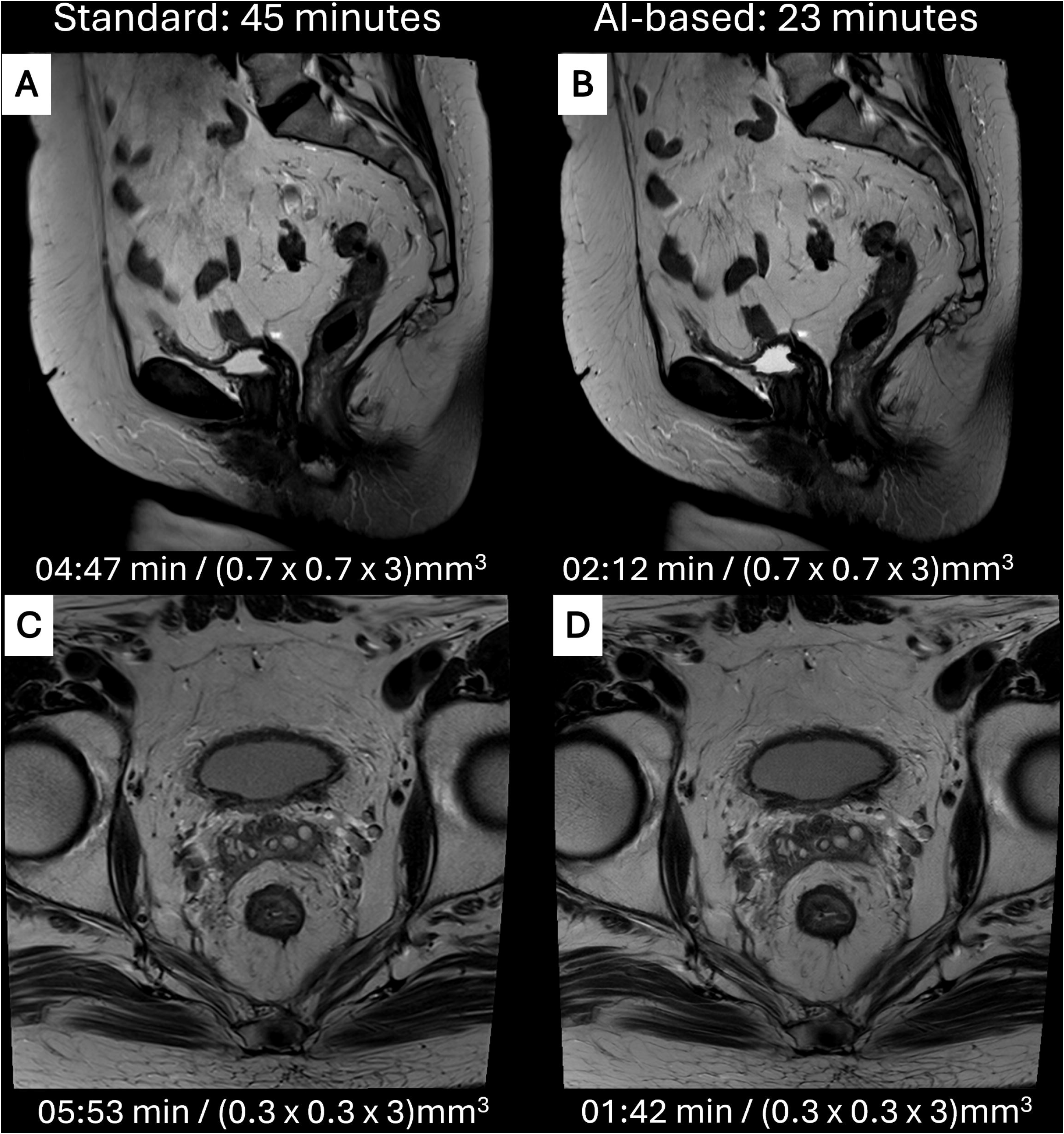
Total average protocol acquisition times, example images, sequence acquisition length and voxel size from the standard protocol (left column) and accelerated protocol using AI-based reconstruction (right column). T2-weighted sagittal TSE images (A,B) and T2-weighted sFOV axial TSE images (C,D).

Deploying the AI-based reconstruction reduced the costs per scan by £47.08. For an institution with similar rates of rectal MRI patients as ours (73 per month, on average), this might hypothetically save £3,436.84 per month. The total cost of the clinical evaluation of the AI-based reconstruction in the rectum protocol was £15,023.23, calculated by considering the cost of 9 volunteer scans plus the total costs of 50 patients scans using both imaging methods minus the total costs of 50 patients scans using standard imaging method (ie,£3,042.23 + £667.69×20 + £617.29×30 - £397.83×50).

### Quality control costing

Table 5 presents the resource use per scan and the total average costs per QC scan, analysed using IQMs. Figure 3 shows a comparison of the cost and acquisition time per scan between each monitoring scheme (ie, no QC, QC using IQMs, QC using radiologist assessment). The total cost per QC scan was £550.84 which is £200.09 more than an AI-based scan without acquisition of QC data due to the involvement of a physicist and additional acquisition time for the QC imaging sequences. A QC scan performed with involvement of a band 8a physicist cost £25.95 more than one with a band 7 physicist. A comparison of monthly savings for the different monitoring schemes is shown in Figure 4. On average, four QC scans were performed per month, which reduced the hypothetical monthly saving to £2,532.68 (ie, £3,436.84 – (£200.09 + £25.95)×4) for a band 8a physicist, or £2,636.48 (ie, £3,436.84 – £200.09×4) for a band 7. physicist. If the IQM assessments were replaced by radiologists’ assessment of image quality which costs £23.70 per scan as estimated in the clinical evaluation phase, the monthly savings would have been reduced by £94.80 considering this additional cost.

**Table 5.**
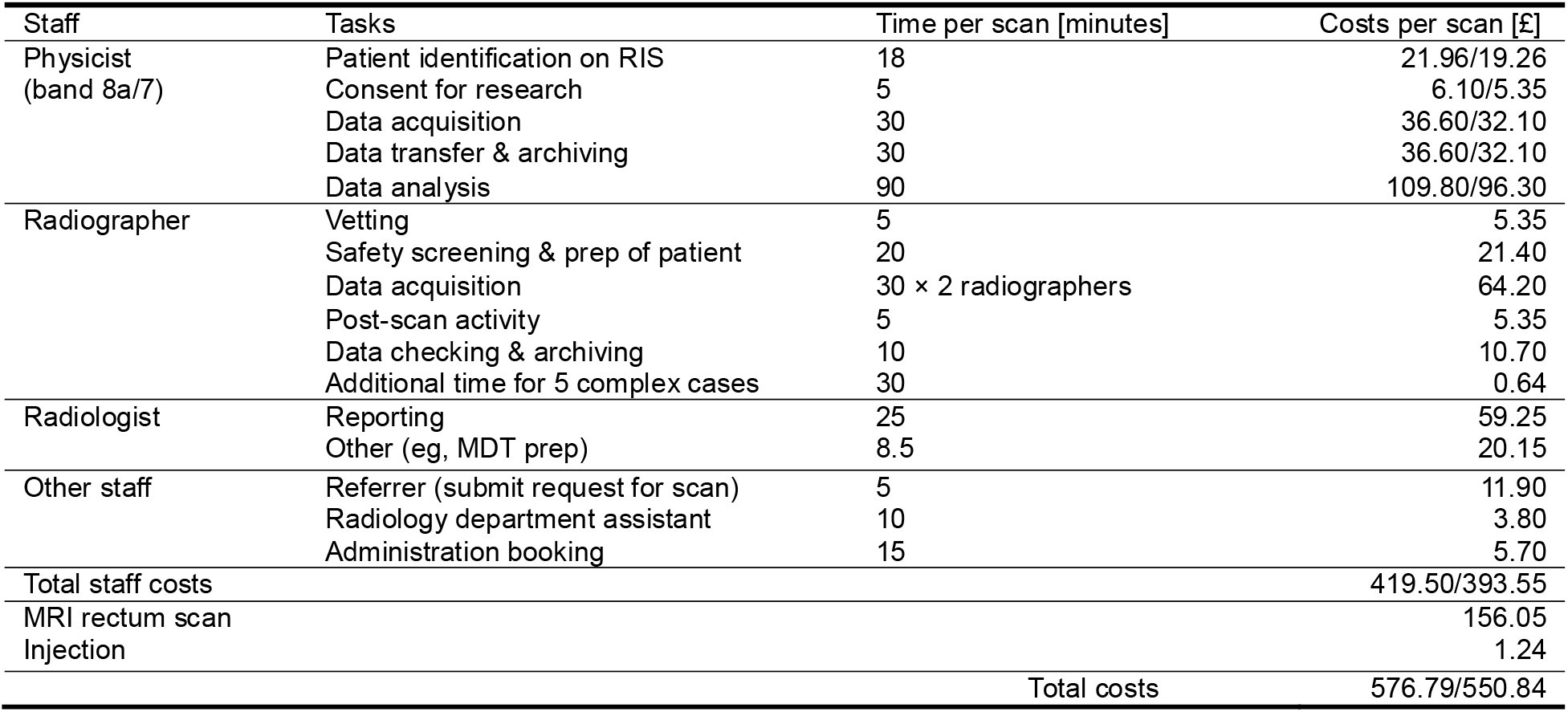
Resource use and costs per QC scan. Costs per scan for physicist tasks and total costs are shown for a band 8a and band 7 physicist.

**Figure 3.**
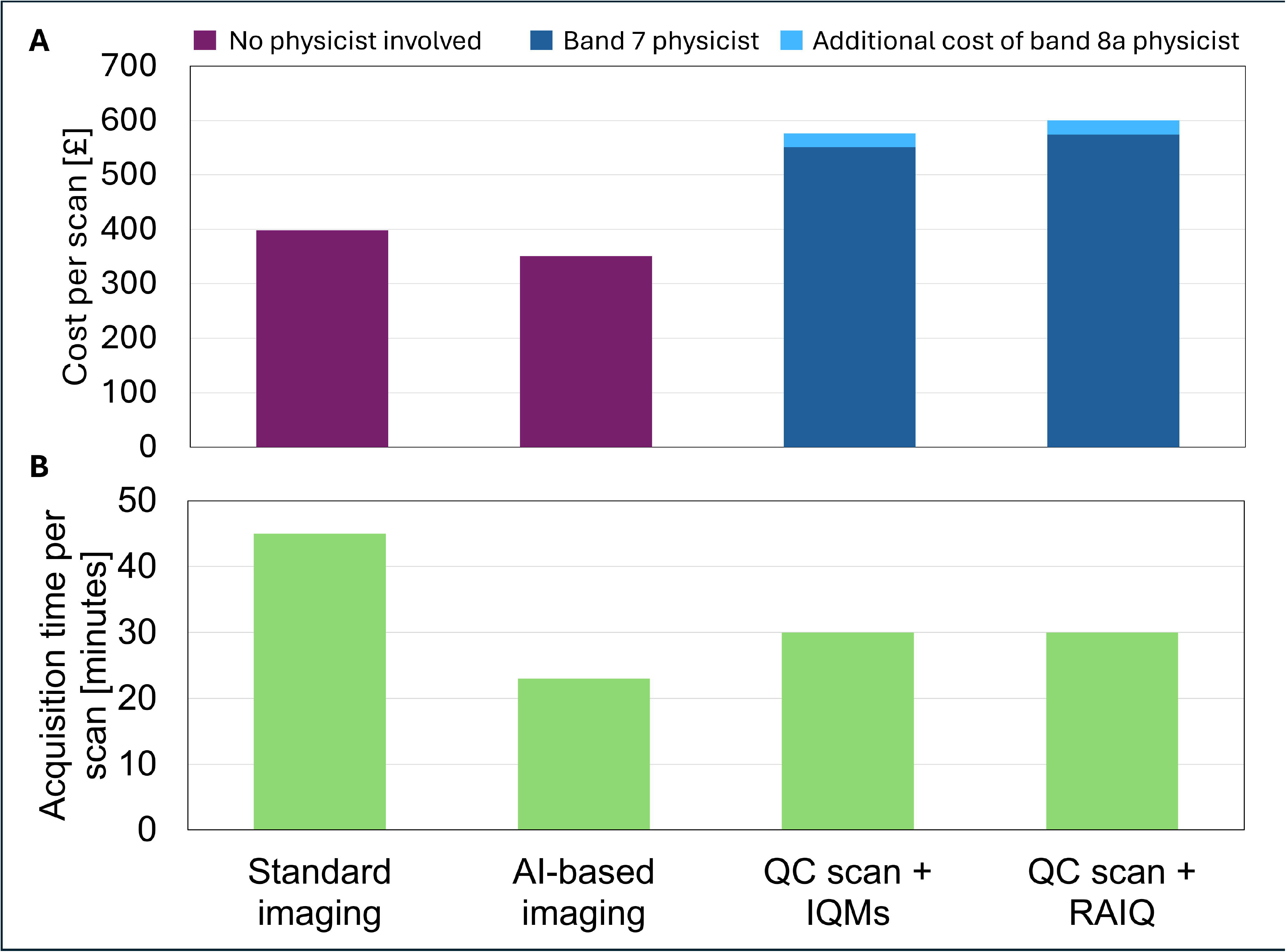
Cost (A) and acquisition time (B) of each type of exam and quality control method. RAIQ = radiologist assessment of image quality.

**Figure 4.**
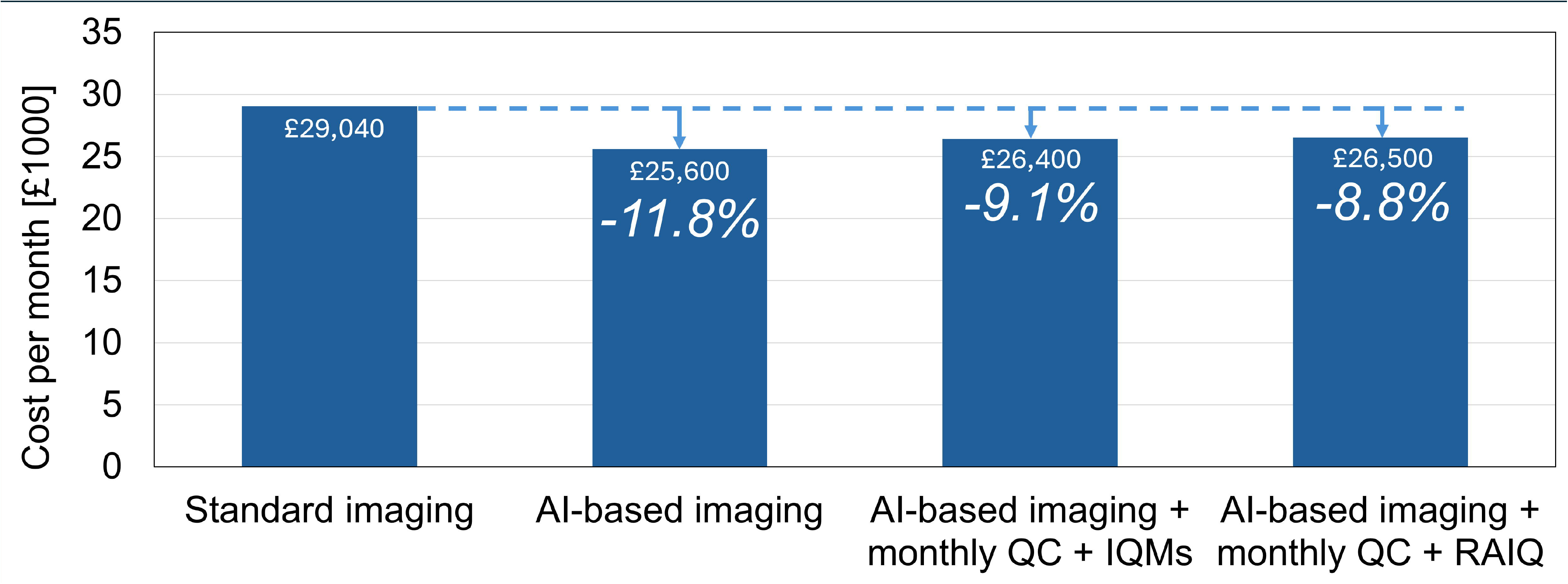
Monthly cost of each type of imaging and quality control method (with a band 7 physicist) with savings relative to standard imaging. Assuming 73 scans per month with 4 QC scans per month (one QC scan per scanner per month).

### Payback period

Estimations of payback periods with each monitoring scheme are shown in Figure 5. The payback period would be 4.4 months for evaluation and deployment of the accelerated protocol with no ongoing quality control. The payback period for accelerating the protocol and performing QC would be 5.8 months for QC using IQMs versus 6.0 months for QC using radiologist assessments considering a band 8a physicist was involved for the first 2 months and a band 7 thereafter.

**Figure 5.**
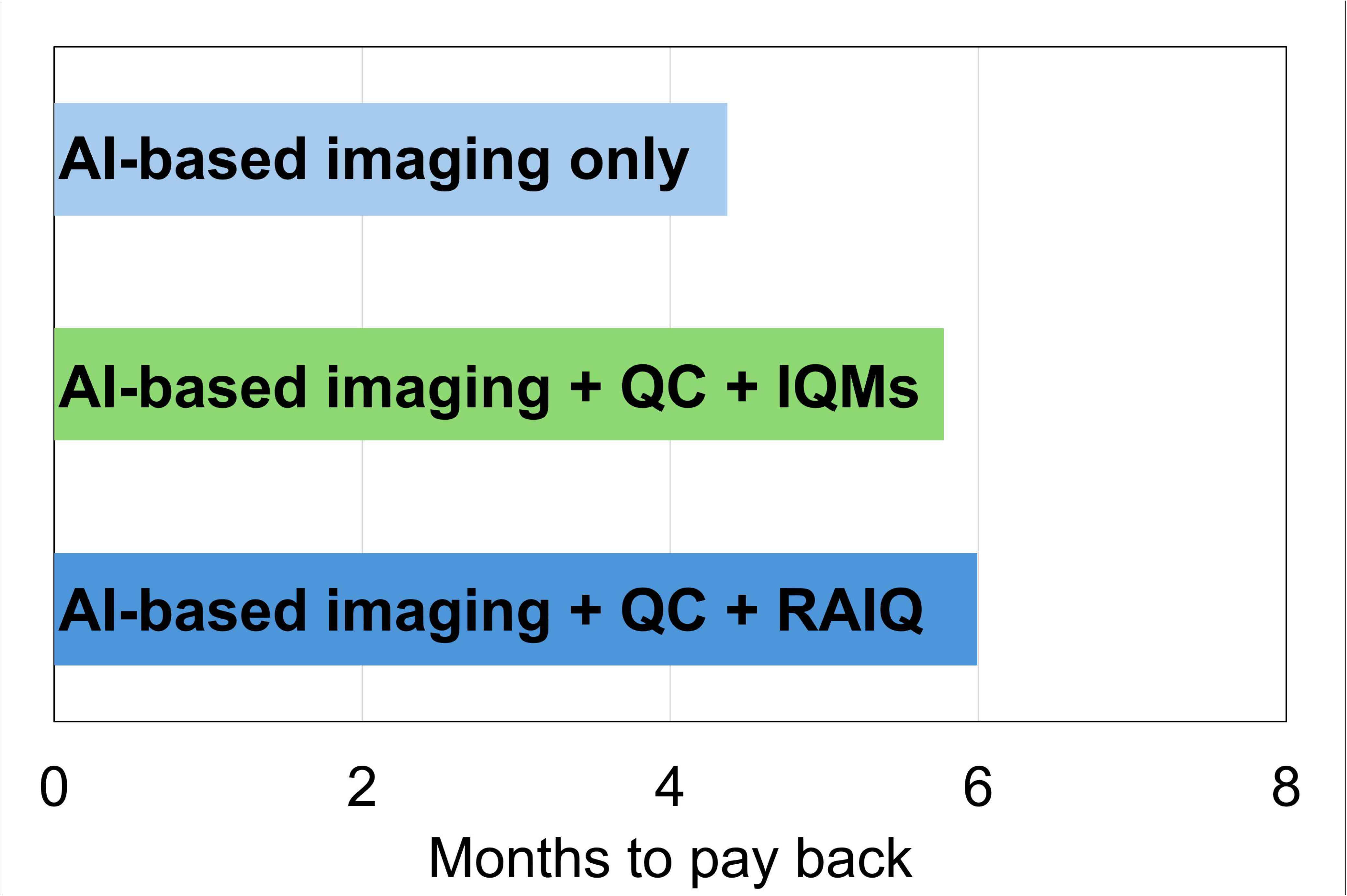
The time taken for the savings of each accelerated imaging and monitoring scheme to pay back the cost of evaluation.

## Discussion

This study evaluated the potential cost savings from implementing an AI-based MR reconstruction to accelerate MRI rectum scans using a simple bottom-up micro-costing method. The evaluation demonstrated the cost of resources required to perform a clinical evaluation and post-deployment monitoring, as recommended by guidelines^2–6^, and the potential costs saved from reduced staff costs per examination via reduced acquisition times.

Tan et al^17^ compared four different resource use costing methodologies, including bottom-up micro-costing, bottom-up gross-costing, top-down micro-costing, and top-down gross costing. However, there is no gold standard in terms of which method is regarded as the best. This study employed a bottom-up micro-costing methodology, given that resources were identified at a very detailed level and the resource use data were collected at the patient level.

It cost our institution £15,023 in resource use to perform nine volunteer scans and 50 patient scans to clinically evaluate an AI-based reconstruction. The cost of the Deep Resolve software licence was unknown and excluded, as it was funded by the NHS England and NHS Improvement (NHSEI) advanced acceleration technology (AAT) fund. Other institutions will need to consider procurement cost of the AI-based AAT. Although the evaluation incurred a substantial cost, it reduced in the average scan length from 45 minutes to 23 minutes, hypothetically reducing costs through reduced radiographer resource use.

A shorter acquisition time also unlocks MRI scanner capacity. An institution could use this to schedule more examinations within the same period, reducing waiting lists or outsourcing scans to the private sector^24^. Alternatively, the capacity could allow more radiographer-patient time, pre- and post-scan, potentially improving patient comfort and experience. Otherwise, capacity could be reserved for urgent inpatient requests. Each institution may decide how to use the additional scanner capacity based on local pressure on the radiology department. For instance, they may need to consider capacity for additional radiological reporting and its associated costs when considering whether to schedule additional scans.

After clinically deploying the accelerated protocol, we chose to perform ongoing monitoring of the AI-based reconstruction by performing QC scans once per scanner per month. Performing each QC scan using IQM analysis cost £200 more than the accelerated scan alone due to additional radiographer resource use for the acquisition and additional physicist resource use for the acquisition and data processing. However, we have shown that using IQMs is a resource-efficient choice compared with assessment of the QC data by radiologists, which would have cost a further £24 per scan.

Our decision to perform the QC scans once per MR system per month was based on the feasibility of data acquisition and processing, the number of clinical accelerated scans and the expected timeframe over which issues may occur. Therefore, despite the added QC cost, accelerating scans with ongoing monitoring still would have saved £2,636 per month compared with standard imaging. These savings would have paid off the £15,023 cost of evaluation after 5.8 months, with net yearly savings of £31,368 thereafter. The benefits from a thorough clinical evaluation and ongoing monitoring scheme are not only financial: investing of resources to implement AI-based reconstructions in the MRI rectum protocol enabled us to reduce the evaluation period when accelerating other MRI protocols. Furthermore, after scanner system upgrades, our QC analysis showed no change the AI-based reconstruction performance^25^.

The estimated cost savings presented in this study reflect the choices we made as an institution. We conducted a rigorous clinical evaluation involving a multidisciplinary team, using substantial numbers of healthy volunteers and patients based on experience, and precedent in the literature. We also developed a resource efficient QC method for ongoing monitoring that did not rely on routine radiological assessment. These choices align with well-established good practices and recommendations in guidelines specific to the use of AI tools in radiology. The guidance lacks consideration of the resources required to follow such recommendations and business cases should consider such resources. This study evaluated the resources required for one NHS trust to implement the recommendations for a single AI tool.

The estimations for times for staff tasks were unique for our institution and the studies evaluated in this work. Radiologist times for reporting may be higher than national averages due to the complexity of the imaging we acquire. However, other institutions can either measure their own reporting times or refer to guidelines for recommended reporting times^26^. Physicist data analysis times may be higher than for routine QC as the times reported account for method development as part of a research project.

This study can serve as a framework for costing the evaluation, deployment and monitoring of other AI-based tool in similar settings. Each implementation step was costed separately so others can selectively replicate the methods, for example using different patient numbers or task timings. Payback time estimates could help a trust prioritise in which clinical protocols it is cost-beneficial to implement AI-based reconstruction. The unit MRI scan cost covers infrastructure, such as scanner procurement and staff training. We did not calculate the total infrastructure cost (eg, capital costs) because the evaluation covered a small percentage of examinations over a short period rather than all examinations a scanner’s lifetime. NHS-level sources were used for the staff resource, MRI rectum scan and antispasmodic injections costs, allowing other NHS trusts to use the same unit costs in their calculations.

This study has limitations. Firstly, we only evaluated one type of imaging examination (MRI rectum) at one institution, however, the cost breakdown facilitates extension to other examinations. Secondly, the evaluation did not include the cost of purchasing the AI tool, however other centres who purchase an AI tool can add this when performing a similar analysis. Thirdly, the estimated cost savings were based solely on reduced of scan times and did not evaluate down-stream resource use, such as additional radiologist reporting, as these would have been hypothetical. Other centres can include this consideration when performing a similar analysis.

## Conclusion

In conclusion, we have evaluated the resource use cost of performing a clinical evaluation and ongoing monitoring of an AI-based MR reconstruction. We found that we would hypothetically pay back the cost of evaluation in 5.8 months from time savings from accelerated acquisitions whilst using resource-efficient IQM analysis to monitor the performance of the reconstruction. This study proposed generalisable methods for other institutions to carry out similar evaluations thus enabling institutions to assess the resources needed to implement an AI-based reconstruction and/or evaluate in which protocols it would be most cost-effective to implement. Future studies could evaluate the resources required to do additional work evaluating the performance of the AI-based reconstruction after MR system software upgrades.

## Data Availability

The data that support the findings of this study are not openly available due to reasons of sensitivity and are available from the corresponding author upon reasonable request. Data are located in controlled access data storage at The Royal Marsden Hospital and the Institute of Cancer Research.

